# Tocilizumab provides dual benefits in treating immune checkpoint inhibitor-associated arthritis and preventing relapse during ICI rechallenge: the TAPIR study

**DOI:** 10.1101/2024.07.12.24310208

**Authors:** Pierre-Florent Petit, Douglas Daoudlarian, Sofiya Latifyan, Hasna Bouchaab, Nuria Mederos, Jacqueline Doms, Karim Abdelhamid, Nabila Ferahta, Lucrezia Mencarelli, Victor Joo, Robin Bartolini, Athina Stravodimou, Keyvan Shabafrouz, Giuseppe Pantaleo, Solange Peters, Michel Obeid

**Affiliations:** Medical oncology service, CHU Helora, Rue Ferrer 159, 7100 La Louvière, Belgium; Centre Hospitalier Universitaire Vaudois (CHUV), University of Lausanne, Department of Medicine, Immunology and Allergy Service, Rue du Bugnon 46, CH-1011 Lausanne, Switzerland; Centre Hospitalier Universitaire Vaudois (CHUV), University of Lausanne, Department of Oncology, Medical Oncology Service, Rue du Bugnon 46, CH-1011 Lausanne, Switzerland

**Author notes:** Corresponding author: Pr Michel Obeid, MD-PhD Lausanne Center for Immuno-Oncology Toxicities LCIT Immunology and Allergy Division, Rue du Bugnon 17, 1011 Lausanne, Switzerland Centre Hospitalier Universitaire Vaudois (CHUV). contributed equally to this work (Pierre-Florent Petit, Douglas Daoudlarian).

**Keywords:** immune checkpoint inhibitors, irAEs, arthritis, secondary prophylaxis, tocilizumab

## Abstract

**Background:** Immune checkpoint inhibitor (ICI)-associated arthritis (ICI-AR) significantly affects quality of life and often requires discontinuation of ICI therapy and initiation of immunosuppressive treatment. The aim of this retrospective study was to evaluate the dual efficacy of tocilizumab (TCZ), an anti-IL-6R agent, in the treatment of ICI-AR and the prevention of relapses after ICI rechallenge.

**Methods:** This retrospective single-center study was conducted at our institution from 2020 to the end of 2023. We identified 26 patients who developed ICI-AR. The primary objectives were to evaluate the therapeutic efficacy of TCZ in the treatment of ICI-AR in 26 patients and to evaluate the potential of TCZ as secondary prophylaxis during ICI rechallenge in 11 of them. For the treatment of ICI-AR, patients received prednisone (CS) at a low dose of 0.3 mg/kg tapered at 0.05 mg/kg weekly for six weeks until discontinuation. TCZ was administered at a dose of 8 mg/kg every two weeks. In the subgroup receiving secondary prophylaxis (rechallenge n=11, in 10 patients), TCZ was reintroduced at the same dosage of 8 mg/kg bi-weekly concurrently with ICI rechallenge, and without the addition of CS. A control group of patients (rechallenge n=5, in 3 patients) was rechallenged without TCZ. Secondary endpoints included post rechallenge evaluation of ICI duration, reintroduction of CS > 0.1 mg/kg/day, ICI-RA flares, and disease control rate (DCR). An additional explanatory endpoint was the identification of biomarkers predictive of response to TCZ.

**Results:** The median age of the patients was 70 years. The median follow-up from ICI initiation was 864 days. ICI regimens included anti-PD-(L)1 monotherapy in 17 patients (63%), anti-PD-1 combined with anti-CTLA4 therapy in 8 patients (31%), and anti-PD-1 combined with anti-LAG3 therapy in 1 patient (4%). Among the 20 patients treated with TCZ for ICI-AR, all (100%) achieved an ACR70 response rate, defined as greater than 70% improvement, at 10 weeks. Additionally, 81% of these patients achieved steroid-free remission after 24 weeks on TCZ.

The median follow-up period was 552 days in rechallenged patients. The ICI rechallenge regimens (n=16) included anti-PD-(L)1 monotherapy in thirteen cases (81%) and combination therapy in three cases (19%). The results demonstrated a reduction in ICI-AR relapses upon ICI rechallenge in patients receiving TCZ prophylaxis as compared to patients who did not receive prophylaxis (17% vs 40%). In addition, the requirement for CS at doses exceeding 0.1 mg/kg/day was completely abolished with prophylaxis (0% vs 20%), and the mean duration of ICI treatment was notably extended from 113 days to 206 days. The 12-month post-rechallenge outcomes showed a disease control rate (DCR) of 77%. Importantly, during TCZ prophylaxis, CXCL9 levels remained elevated, showing no decline from their levels at the onset of ICI-AR. Additionally, elevations of IL-6 and CXCL10 levels were exclusively observed in patients who developed new irAEs during the period of TCZ prophylaxis.

**Conclusion:** In addition to its efficacy in treating ICI-AR, TCZ demonstrated efficacy as a secondary prophylactic agent, preventing the recurrence of ICI-AR symptoms and lengthening ICI treatment duration after ICI rechallenge. The use of TCZ as a secondary prophylaxis may represent a promising strategy to extend patient exposure to ICI treatments and maximize therapeutic benefit.

**Highlights:**

1- TCZ achieved a 100% ACR70 response rate at 24 weeks, demonstrating its efficacy in the treatment of ICI-RA.
2- A significant 81% of patients achieved steroid-free status after 24 weeks on TCZ, underscoring its utility in accelerating CS tapering.
3- TCZ serves as a successful secondary prophylaxis in patients rechallenged with ICI, preventing significant arthritis flares and the need for additional CS use.
4- TCZ prophylaxis reduces the median time to ICI rechallenge by 47.5 days and extends the duration of uninterrupted ICI therapy by 93 days.
5- CXCL9 levels were not reduced during TCZ prophylaxis, suggesting that there was no negative impact on cytokines associated with oncologic response. In addition, early increases in IL-6 and CXCL10 levels may signal the onset of new irAEs during prophylaxis.

## Introduction

Immune checkpoint inhibitor-associated arthritis (ICI-AR) can be persistent and significantly impacts patients’ quality of life, necessitating discontinuation of ICI therapy and long-term use of immunosuppressants^1, 2^. Although clinical trials report arthralgia incidence rates ranging from 5 to 11%, expert opinion suggests that these immune-related adverse events (irAEs) may be underreported^3, 4^. The most common rheumatic (rh) rh-irAEs include inflammatory polyarthritis and polymyalgia rheumatica (PMR)-like syndromes^5–7^. In contrast to the classical findings observed in patients with rheumatoid arthritis, patients with ICI-AR are rarely positive for rheumatoid factor (RF) or anti-citrullinated protein antibodies (ACPA)^4^. The pathogenesis of ICI-RA remains incompletely understood. The composition of immune cells in the synovial fluid differs significantly from that observed in patients with rheumatoid or psoriatic arthritis^8^. Recent studies have identified a significant clonal expansion of activated effector CD8^+^ T cell populations, including CD38^hi^CD127^-^ and CXCR3^hi^ subsets in synovial fluid, and CX3CR1^hi^ CD8^+^ T cells in blood^9, 10^. In contrast to IL-6, levels of IFN-γ, TNF-α, and the Th17 signature are reduced in both peripheral blood and synovial fluid following immunosuppressive treatment in ICI-RA patients^9^. However, the precise extent of inflammatory cytokine fluctuation relative to baseline levels in cancer patients prior to ICI treatment remains unknown, representing a substantial gap in our data continuum. The lack of comprehensive data on the fluctuations of cytokines limits the identification of reliable biomarkers for the monitoring of ICI-RA progression and the prediction of treatment outcomes. Consequently, the establishment of comprehensive baseline and longitudinal cytokines profiling is of paramount importance for the advancement of our understanding and the improvement of ICI-RA management

Tocilizumab (TCZ), an interleukin-6 receptor (IL-6R) inhibitor, is approved for the treatment of many rheumatic diseases, including rheumatoid arthritis, systemic juvenile idiopathic arthritis and giant cell arteritis. It has recently emerged as a promising therapeutic alternative for the treatment of irAEs due to its specificity and potential to attenuate systemic inflammation without suppressing T-cell activity or compromising the anti-tumor immune response^11–13^. Recent studies have demonstrated the efficacy of TCZ in the treatment of rheumatoid irAEs while preserving the anti-tumor immune response^14–17^.

Furthermore, the incidence of identical irAEs flares following ICI rechallenge exhibits considerable variability, with reported recurrence rates ranging from 18% to 42% across various studies^18–22^. These relapses can be of increased severity, particularly if the initial irAEs were of high grade. Notwithstanding the difficulties inherent to ICI therapy resumption following severe irAEs, it remains a viable option for a subset of patients who have previously demonstrated a favorable response to ICI retreatment. Our team has previously proposed the use of selective immunosuppression (SI), such as TCZ for arthritis and vedolizumab for colitis, as a secondary prevention when restarting ICI treatment^22^. Previous reports on prophylaxis for ICI-associated colitis have indicated that the recurrence rate of severe ICI-associated colitis following ICI rechallenge was lower in patients receiving prophylaxis with vedolizumab or infliximab compared to those not receiving prophylaxis^23, 24^.These preventive approached aim to achieve a balance between efficacy and safety, taking advantage of the favorable safety profile of SI to facilitate ICI retreatment. Nevertheless, the efficacy of TCZ as a secondary prophylaxis in the reintroduction of ICI therapy after IRI-RA has not been subjected to a comprehensive evaluation.

The primary objective of this study is to investigate the dual functionality of TCZ as a treatment of ICI-AR and as a prophylactic agent during subsequent ICI rechallenge. A retrospective analysis was conducted on a cohort of 26 cancer patients diagnosed with ICI- AR. This analysis included sequential deep inflammatory profiling of 57 cytokines at four critical time points: baseline prior to the initiation of ICI therapy, at the time of ICI-AR diagnosis, prior to ICI rechallenge, and during ICI rechallenge with TCZ prophylaxis. The data from these patients were compared with those from a larger cohort of cancer patients prior to ICI treatment (n=164) to ascertain inflammatory profiles associated with the onset of ICI-AR and to identify biomarkers predictive of the efficacy of TCZ, both as a treatment and as a prophylactic measure.

## RESULTS

### Clinical demographic and rh-irAE characteristics

A total of 31 patients treated at the Lausanne University Hospital between 2020 and the end of 2023 were identified with rh-irAEs following ICI treatment. Of these, 26 patients with ICI-related arthritis (ICI-RA) were included in the final analysis, as described in the study flowchart (**Supplementary Figure 1**). Patient characteristics are summarized in Table 1. The mean age of the patients was 70 years, with a male predominance (n=15/26, 58%). The most common types of cancer were melanoma and lung cancer, with 11 out of 26 patients (42%) for each tumor type. The majority of the patients were treated with anti-PD-1 monotherapy (n=16/26, 62%) or the combination of nivolumab plus ipilimumab (n=8/26, 31%).

**Table 1.** Clinical, demographic, biological and rh-irAEs characteristics of the ICI-AR cohort. The principal clinical, biological, demographic, and rh-irAEs characteristics for the ICI-AR cohort (n=26) are detailed in this table.

|  | n = 26 | % within cohort |
| --- | --- | --- |
| **<u>Age</u>** |  |  |
| Mean at rh-irAE onset | 70 | 95% CI:63-77 |
| **<u>Cohort follow-up (median, days)</u>** |  |  |
| Follow-up from ICI initiation | 864.5 | 95% CI:576-1152 |
| Follow-up from rh-irAE | 564 | 95% CI: 384-825 |
| **<u>rh-irAEs onset</u>** |  |  |
| Median time of onset (days) | 171 | 95% CI:53-326 |
| Onset within 90 days | 11 | 42% |
| **<u>rh-irAEs clinical presentation</u>** |  |  |
| Polyarticular arthritis (>5 joints) | 14 | 54% |
| PMR-like | 7 | 27% |
| Oligo-arthritis (<5 joints) | 5 | 19% |
| **<u>Rh-irAE duration (median, days)</u>** |  |  |
| Rh-irAE duration | 360 | 95% CI:271-546 |
| **<u>Biological findings</u>** |  |  |
| **<u>Rheumatoid factor (RF)</u>** |  |  |
| Tested positive | 0/26 | 0% |
| **<u>ACPA</u>** |  |  |
| Tested positive | 0/26 | 0% |
| **<u>Gender</u>** |  |  |
| Male | 15 | 58% |
| Female | 11 | 42% |
| **<u>Tumor type</u>** |  |  |
| Melanoma | 11 | 42% |
| Lung | 11 | 42% |
| Head and neck | 2 | 8% |
| Colorectal | 1 | 4% |
| Renal cell carcinoma | 1 | 4% |
| **<u>Initial ICI therapy</u>** |  |  |
| Nivolumab/ipilimumab | 8 | 31% |
| Nivolumab/relatlimab | 1 | 4% |
| Anti PD-1 monotherapy | 16 | 62% |
| Anti PD-L1 monotherapy | 1 | 4% |
| **<u>Pre-existing rheumatic diseases</u>** |  |  |
| Polymyalgia rheumatica | 3 | 12% |
| Seronegative polyarthritis | 1 | 4% |
| Axial spondyloarthritis | 1 | 4% |
| **<u>Concomitant irAE</u>** |  |  |
| Colitis | 3 | 12% |
| Cutaneous | 2 | 8% |
| Hepatitis | 2 | 8% |
| Endocrine | 4 | 15% |
| Pneumonitis | 1 | 4% |
| Myositis | 2 | 8% |

The median time from the onset of ICI treatment to the onset of rh-irAEs was 171 days (95% CI: 53-326). The median follow-up period following the initial ICI administration was 864 days (95% CI: 576-1152). The median duration of rh-irAEs was 360 days (95% CI:271-546). The most common clinical presentation was polyarticular arthritis (n=14, 54%), while PMR- like arthralgia (n=7, 27%) and oligoarticular arthritis (n=5, 19%) were less prevalent. All patients tested negative for rheumatoid factor (RF) and anti-citrullinated protein antibodies (ACPA) (n=0/26, 0%). A history of pre-existing non-active inflammatory rheumatic disease was documented in only 20% (n=5/26) of patients, and none were receiving immunosuppressive drugs (IS) at the time of ICI initiation. A proportion of patients (n=17/26, 53%) developed other concomitant immune-related adverse events (irAEs) (**Table 1**).

### Tocilizumab is an effective treatment for ICI-AR and allows CS tapering

In the patient group (n=20/26, 77%) treated with biweekly TCZ at a dosage of 8 mg/kg, in combination with a low dose of prednisone CS at 0.3 mg/kg/day, 100% of the patients achieved an ACR70 arthritis score^25–27^ (indicating at least 70% improvement from baseline) by weeks 10-12. Notably, this rapid improvement was also observed in two patients who received TCZ without CS (n=2/20, 10%). TCZ consistently enabled patients to achieve ACR70 score at both 10-12 and 24-weeks following the onset of ICI-AR, as shown in **Table 2**. 24 weeks (24W) after the introduction of TCZ, 81% of patients (n=13/16) achieved a steroid-free status. The mean CS dose was 23 mg and the average time to reduce CS to less than 10 mg after starting TCZ was 30.7 days. The median duration of TCZ treatment was 235 days (95% CI: 55-450). In contrast, those treated with CS alone (n=6) had less favorable outcomes. At 24 weeks, only 25% of these patients achieved steroid-free status, compared to 81% in the TCZ-treated group. In addition, this group required a higher mean CS dose of 37.5 mg compared to 23 mg in the TCZ group, and they required longer time - an average of 40.25 days versus 30.7 days - to taper their CS dose to below 10 mg (**Table 2**). This comparison underscores TCZ rapid efficacy in treating ICI-AR and highlights the associated benefits in terms of reducing both CS dosage and treatment duration.

**Table 2.**
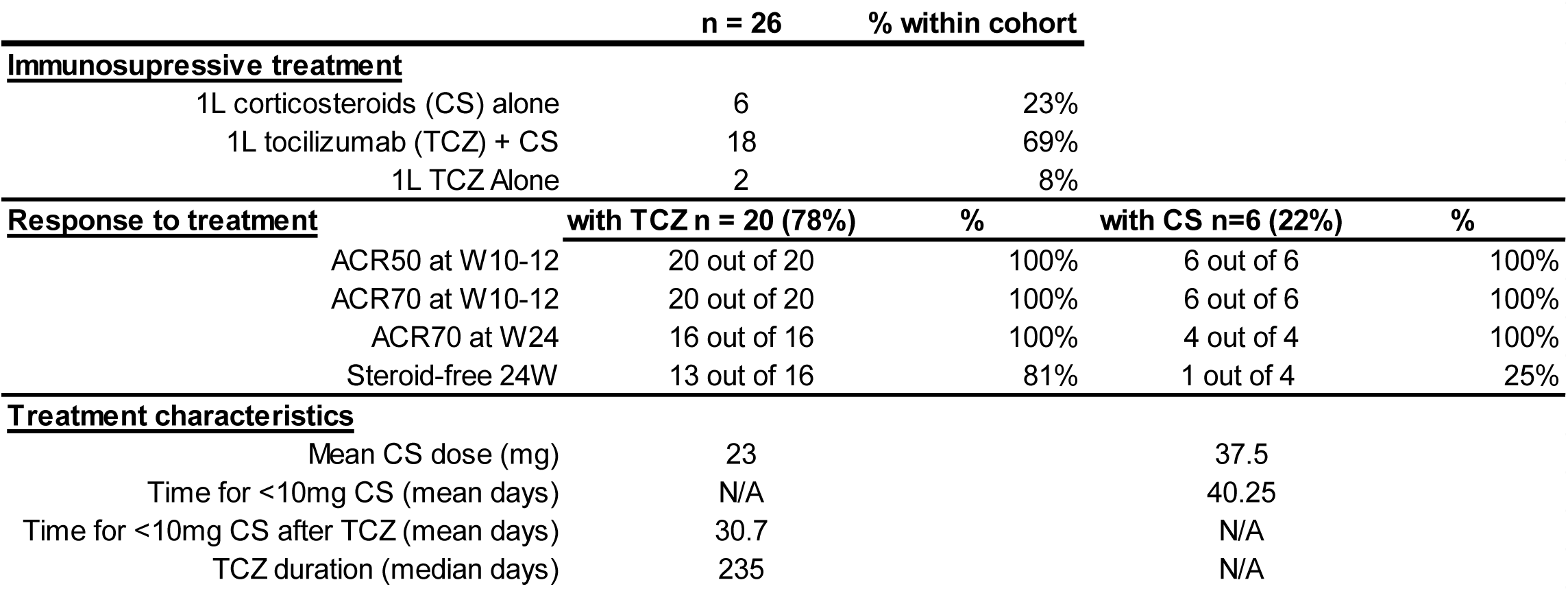
Immunosuppressive treatments and patient responses in the ICI-AR cohort. The different types of immunosuppressive treatment, and symptoms outcomes under treatment in the ICI-AR cohort (n=26) are detailed in this table. 6 patients were treated with CS alone, 18 patients were treated with a combination of TCZ 8 mg/kg Q2w + low dose CS at 0.3 mg/kg/day and 2 patients were treated with TCZ 8mg/kg Q2W alone without CS. The ACR50/70 at 10-12w and 24w, the steroid-free status at 24w are detailed. The mean dose of CS and the mean time for <10mg CS in each treatment group are detailed. CS, corticosteroids; TCZ tocilizumab.

### Tocilizumab prophylaxis facilitates ICI rechallenge and reduces ICI-AR relapse

After completing CS tapering and the resolution of ICI-AR, 13 patients underwent ICI rechallenge, including three undergoing rechallenge twice, for a total of 16 rechallenge episodes (RCLs). TCZ was employed as secondary prophylaxis (TP) in 11 RCLs, while the remaining 5 RCLs proceeded without TCZ prophylaxis (WP). The same ICI was reintroduced in 91% of RCLs (n=10/11) in the TP subgroup, compared to 60% (n=3/5) in the WP subgroup. The median interval between end of initial ICI and ICI rechallenge was 70 days (95% CI:56-84) in the WP group vs 22.5 days (95% CI:20-295) in the TP group, with a median follow-up duration post-rechallenge of 498 days (95% CI:341-1040) in the TP group vs 566 days (95% CI:129-757) in the WP group. At the time of ICI rechallenge, oncological disease was progressing in 6 cases—40% in the WP subgroup and 36% in the TP subgroup.

In the TP subgroup (n=11), the incidence of ICI-AR relapses decreased to 18%, compared to 40% in the WP group. Moreover, the requirement for reintroduction of CS at doses exceeding 0.1 mg/kg/day dropped to 0% in the TP group versus 20% in the WP group. The mean duration of ICI treatment in the TP group notably extended to 206 days (95% CI:78-333) compared to 113 days (95% CI:16-210) in the WP group. At the last follow-up, ongoing ICI treatment was observed in 36% (n=4/11) of the TP group, compared to none in the WP group. In the TP group, the percentage of ICI interruption post-rechallenge due to irAEs was 27%, as opposed to 40% in the WP group. These results are summarized in Table 3.

**Table 3.** Efficacy of secondary prophylaxis with TCZ in rechallenged patients. . The main clinical characteristics and treatment response of the two groups are described, the TCZ secondary prophylaxis (TP) (Rechallenge n=11, in 10 patients) compared to the rechallenged group without TCZ prophylaxis (WP) (Rechallenge n=5, in 3 patients). The median interval between end of initial ICI and rechallenge, the recurrence of rh-irAEs ≥G2, the need for CS>0.1mg/kg/day, the mean duration of ICI treatment and the reason for ICI discontinuation are detailed. PD; progressive disease, G1 and G2; grade 1 and 2, CRS; cytokine release syndrome, CI confidence interval.

|  |  | Without TCZ Prophylaxis |  | With TCZ Prophylaxis |  |
| --- | --- | --- | --- | --- | --- |
|  |  | n = 5 | % | n = 11 | % |
| Patients with >1 rechallenge |  | 2 | 40% | 1 | 9% |
| Median Interval between end of initial ICI and rechallenge |  | 70 | 95% CI:56-84 | 22.5 | 95% CI:20-295 |
| Rechallenge for PD |  | 2 | 40% | 4 | 36% |
| **<u>Tumor type</u>** |  |  |  |  |  |
| Lung |  | 2 | 67% | 5 | 50% |
| Melanoma |  | 1 | 33% | 4 | 40% |
| Colorectal |  | 0 | 0% | 1 | 10% |
| **<u>Rechallenge as compared to initial ICI therapy</u>** |  |  |  |  |  |
| Similar ICI |  | 3 | 60% | 10 | 91% |
| Nivolumab to ipilimumab/nivolumab |  | 0 | 0% | 1 | 9% |
| Nivolumab to pembrolizumab/chemotherapy |  | 1 | 20% | 0 | 0% |
| Nivolumab to nivolumab/relatlimab |  | 1 | 20% | 0 | 0% |
| **<u>rh-irAE evolution</u>** |  |  |  |  |  |
| Requiring steroids >0.1 mg/kg for Arthritis |  | 1 | 20% | 0 | 0% |
| Relapse rh-irAE ≥ G2 |  | 2 | 40% | 2 | 18% |
| **<u>Other irAEs</u>** |  |  |  |  |  |
| Colitis ≥ G2 |  | 1 | 20% | 5 | 45% |
| CRS G1 |  | 0 | 0% | 1 | 9% |
| Pneumonitis |  | 1 | 20% | 0 | 0% |
| **<u>ICI rechallenge interruption</u>** |  |  |  |  |  |
| Due to rh-irAEs flares |  | 0 | 0% | 0 | 0% |
| Due to others irAEs types |  | 2 | 40% | 3 | 27% |
| Due to disease control |  | 1 | 20% | 1 | 9% |
| Due to neutropenia |  | 0 | 0% | 1 | 9% |
| Due to end of adjuvant treatment |  | 0 | 0% | 1 | 9% |
| Due to PD |  | 2 | 40% | 0 | 0% |
| Patient decision |  | 0 | 0% | 1 | 9% |
| **<u>ICI treatment duration</u>** |  |  |  |  |  |
| Mean duration (days) |  | 113 | 95% CI:16-210 | 206 | 95% CI:78-333 |
| ICI > 90 days |  | 2 | 40% | 6 | 55% |
| Ongoing ICI at least follow-up |  | 0 | 0% | 4 | 36% |
| Median follow up after rechallenge (days) |  | 566 | 95% CI:129-757 | 498 | 95% CI:341-1040 |

These results indicate that TCZ prophylaxis can reduce the recurrence of ICI-AR and prevent the escalation of CS use. In addition, TCZ was associated with a shorter interval between initial ICI cessation and ICI rechallenge, and with lower incidence of new irAE development. Consequently, TCZ prophylaxis prolonged the duration of ICI treatment.

### Oncological outcome and treatment response

Table 4 describes the oncologic outcome after ICI-RA in patients with definitive ICI discontinuation (n=13). In these patients, the last ICI injection was administered on average 19.6 days after the start of ICI-RA. We observed few cases of progressive disease in these patients (n=1, 8%), while some presented oligo-metastatic progression that was treated with local therapy (at 6 months, n=3; at 12 months, n=2). Disease control rates (DCR) were 78% at 6 months and 71% at 12 months in the absence of systemic treatment. We then evaluated the oncological response in rechallenged patients. These patients had worse outcomes after ICI-RA initiation, as 4 patients were rechallenged due to disease progression. Nevertheless, the DCR after rechallenge was 82% at 6 months and 70% at 12 months. The swimmer plot in Figure 1 illustrates ICI treatment and oncological outcomes of patients over time.

**Figure 1.**
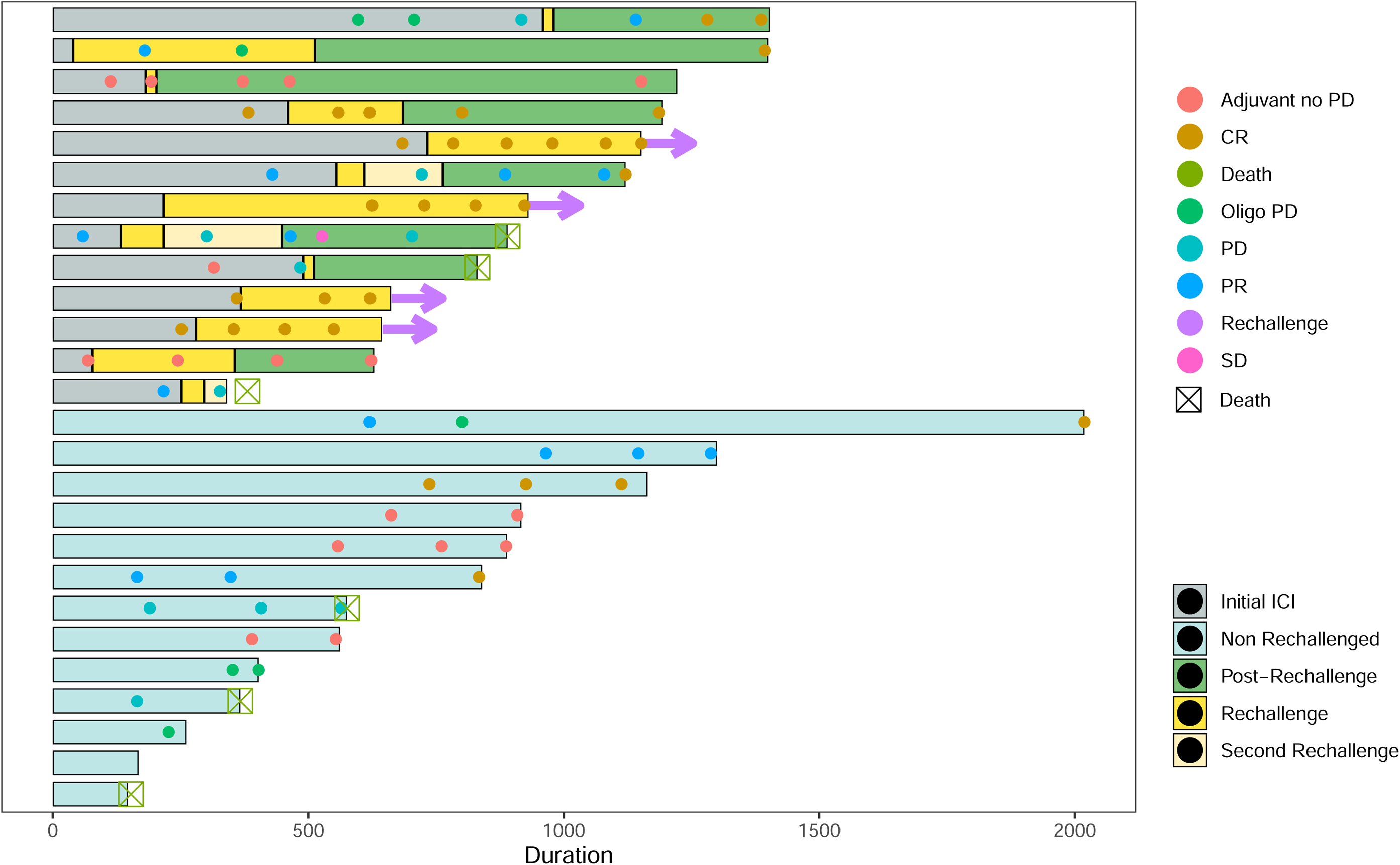
Swimmer plot showing duration of oncological response in relationship to duration of ICI treatment and time of ICI rechallenge in the non-rechallenged (n=13) and rechallenged (n=13) groups. Individual swimmer plots for each patient (n=26). CR, complete response, PR, partial response, PD progressive disease, Oligo-PD, oligo-progressive disease. Arrows indicate in progress ICI rechallenge treatment.

Due to the retrospective design of the study, the 2 cohorts (no rechallenge and rechallenge) were biased by oncologist decisions. Patients who develop ICI-RA are commonly associated with a good prognosis. In our cohort, ICI rechallenge was preferentially performed in patients with worse oncologic prognosis, with 4 patients out of 13 patients presenting disease progression at the time of rechallenge initiation (**supplementary Table 1**).

ICI rechallenge was performed with TCZ prophylaxis in 10 of the 13 patients. Oncologic responses were likely achieved due prolonged exposure to ICI treatment. These results underscore the potential of TCZ prophylaxis not only for the management and prevention of ICI-RA, but also for the prolongation of ICI treatment leading to durable disease control.

### A comprehensive profile of cytokine and biological variation

To investigate the inflammatory profile associated with ICI-AR, we evaluated a large spectrum of classical and inflammatory biomarkers prior to the introduction of immunosuppressive therapy. Our analysis focused on the serum levels of 57 cytokines, chemokines, and growth factors. Among these, only four—CXCL9, CXCL10, IL-6, and the anti-inflammatory cytokine IL-1RA— demonstrated elevated levels at the onset of ICI-AR compared to the baseline levels in cancer patients prior to starting ICI therapy (**Figure 2 and supplementary table 2**). At the time of rechallenge, after the cessation of CS and TCZ treatment, the levels of CXCL9, CXCL10, and IL-1RA continued to be elevated while levels of IL-6 did not. This pattern suggests a sustained inflammatory response independent of IL-6.

**Figure 2.**
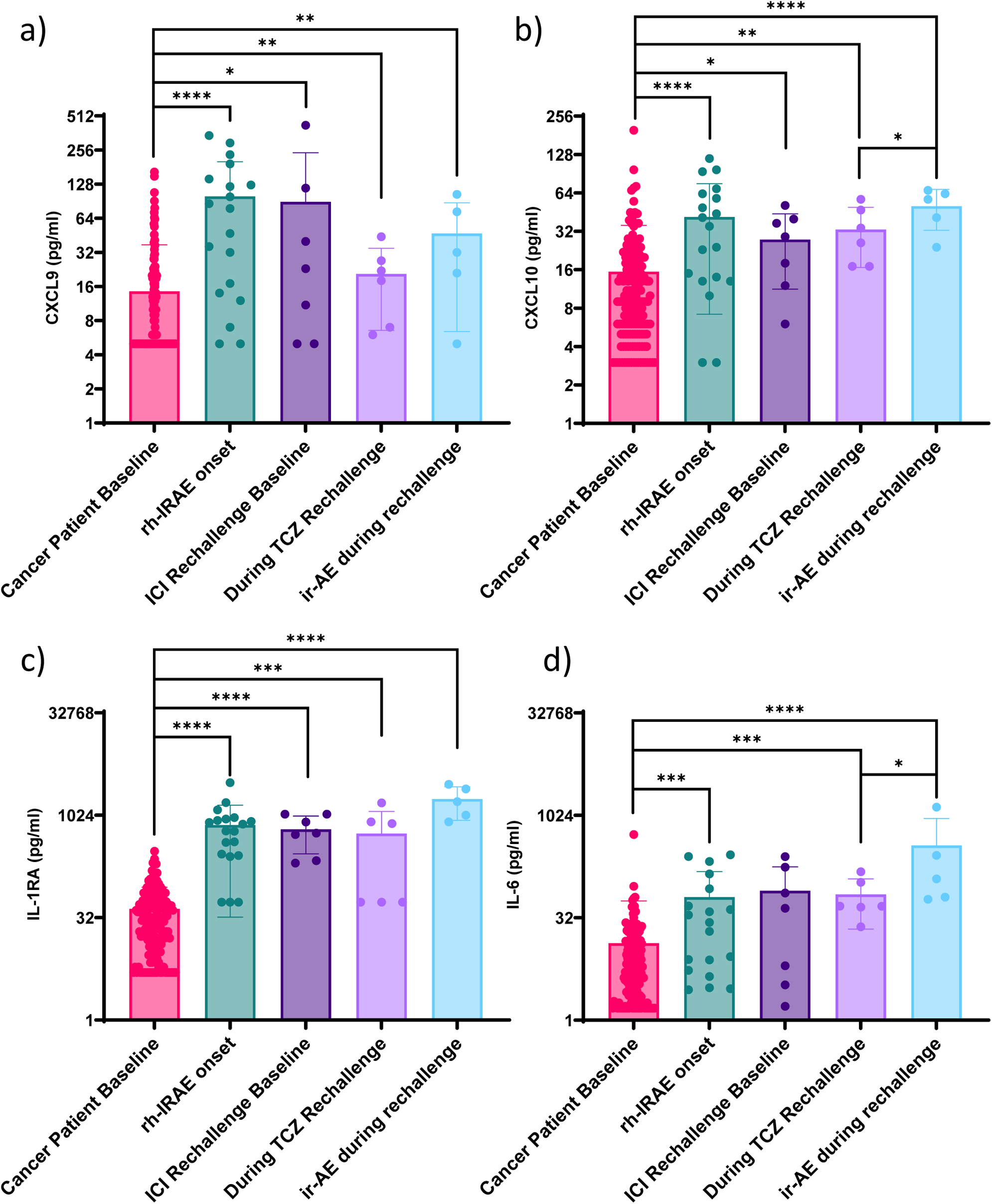
Comprehensive cytokine and biological profiles at different timepoints compared to cancer patient baseline. (**a-f**), Evaluation of serum biomarkers in ICI-AR patients at four time points: the time of ICI-RA onset (n=19), ICI rechallenge baseline (n=7), during ICI rechallenge with TCZ prophylaxis (n=6) and at the time of new irAEs during ICI rechallenge under TCZ prophylaxis (n=5) compared to the pre-ICI serum levels in cancer patients before ICI treatment (n=164) used as the reference group. (a-d) serum levels of chemokines, growth factors and cytokines during the four time points. Mann-Whitney U test tests were used to analyze the data for statistical significance between groups. Plots represent values with individual data points, bar represent the mean and error bars represent standard deviation. The results showed a significant difference between the groups with a P-value of less than 0.05 (*), less than 0.01 (**), less than 0.001 (***) and less than 0.0001 (****).

During ICI rechallenge with TCZ prophylaxis, the levels of these cytokines were relatively stable, consistent with their levels at ICI-AR onset and at RCL baseline. Notably, the levels of CXCL10 and IL-6 increased upon emergence of new irAEs following ICI rechallenge under TCZ prophylaxis, indicating their potential utility in predicting the development of new irAEs despite ongoing TCZ prophylaxis (**Figure 2 and supplementary table 2**).

## Figures legend

**Supplementary Figure 1:**
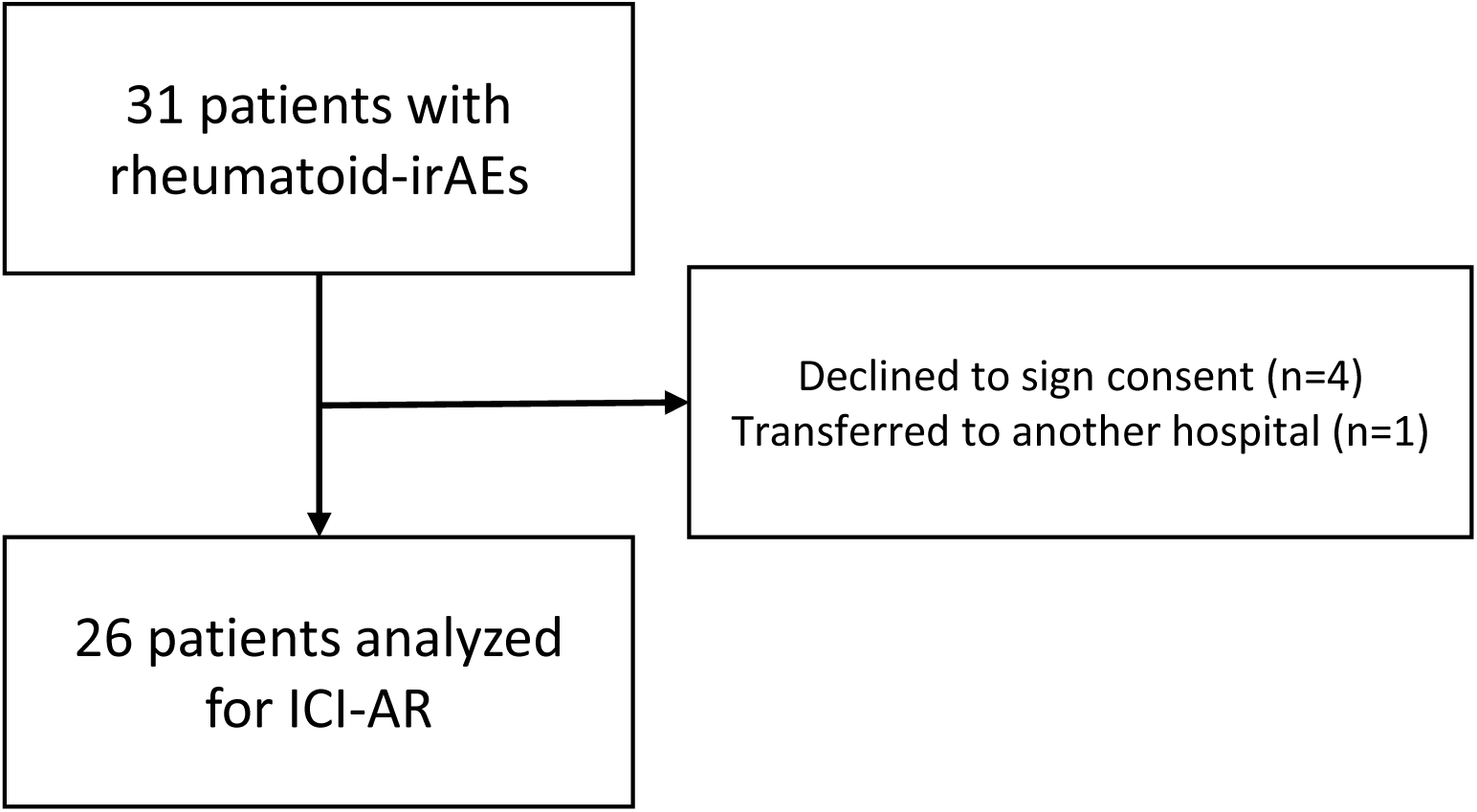
Study flowchart.

**Supplementary Figure 2:**
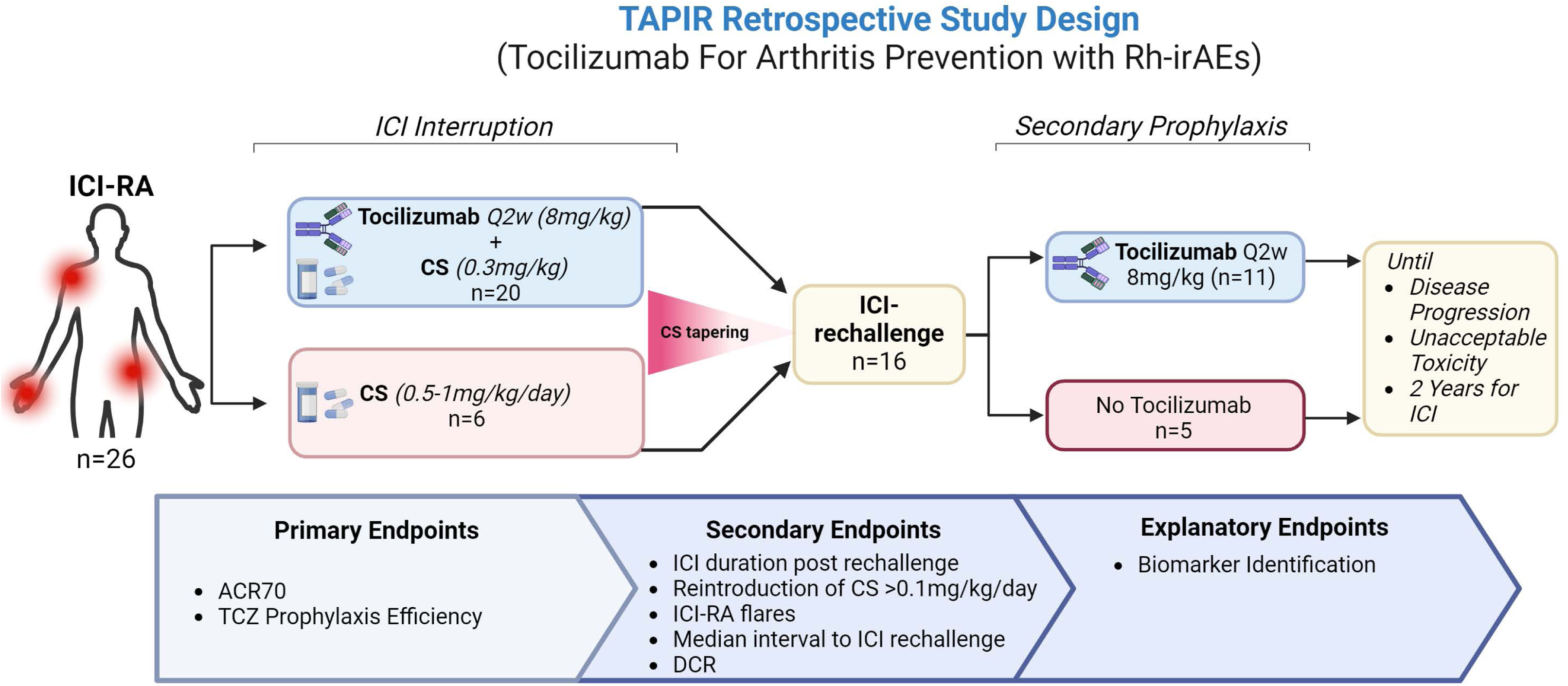
TAPIR retrospective study design, Tocilizumab For Arthritis Prevention with Rh-irAEs.

**Supplementary Table 1.** Oncological outcomes at 6 and 12 months after initial ICI-RA in non-rechallenged (n=13) and rechallenged population (n=13), and after ICI rechallenge in the rechallenged group. The objective response rate (ORR) and disease control rate (DCR) are detailed in the group of patients without ICI rechallenge (n=13) and in the group of patients with ICI rechallenge (n=12, 11 patients on TCZ prophylaxis) 6 and 12 months after the initial ICI-RA. The lower part of the table details disease control rate (DCR) at 6 months and 12 months from rechallenge in the rechallenged group.

|  | Patient without ICI<br>rechallenge (n=13) |  | Patient with ICI<br>rechallenge (n=13) |  |
| --- | --- | --- | --- | --- |
| **<u>Oncological outcome 6 months after initial rh-irAE</u>** |  |  |  |  |
| Complete Response (CR) | 1 | 8% | 3 | 23% |
| Partial Response (PR) | 3 | 23% | 3 | 23% |
| Adjuvant no PD | 3 | 23% | 2 | 15% |
| Oligo-PD | 3 | 23% | 2 | 15% |
| PD | 1 | 8% | 3 | 23% |
| Death | 1 | 8% | 0 | 0% |
| Unknown | 1 | 8% | 0 | 0% |
| Disease Control Rate (DCR) | 7 | 78% | 8 | 73% |
| **<u>Oncological outcome 12 months after initial rh-irAE</u>** |  |  |  |  |
| Complete Response (CR) | 1 | 8% | 4 | 31% |
| Partial Response (PR) | 2 | 15% | 2 | 15% |
| Adjuvant no PD | 3 | 23% | 2 | 15% |
| Oligo-PD | 2 | 15% | 2 | 15% |
| PD | 0 | 0% | 2 | 15% |
| Death | 2 | 15% | 1 | 8% |
| Unknown | 3 | 23% | 0 | 0% |
| Disease Control Rate (DCR) | 5 | 71% | 8 | 73% |
| **<u>Oncological outcome 6 months after ICI rechallenge</u>** |  |  | **n=11** |  |
| Complete Response (CR) |  |  | 4 | 36% |
| Partial Response (PR) |  |  | 3 | 27% |
| Stable Disease (SD) |  |  | 2 | 18% |
| Progressive Disease (PD) |  |  | 1 | 9% |
| Death |  |  | 1 | 9% |
| Disease Control Rate (DCR) |  |  | 9 | 82% |
| **<u>Oncological outcome 12 months after ICI rechallenge</u>** |  |  | **n=10** |  |
| Complete Response (CR) |  |  | 6 | 60% |
| Partial Response (PR) |  |  | 1 | 10% |
| Stable Disease (SD) |  |  | 0 | 0% |
| Progressive Disease (PD) |  |  | 2 | 20% |
| Death |  |  | 1 | 10% |
| Disease Control Rate (DCR) |  |  | 7 | 70% |

**Supplementary Table 2. Global cytokine table.** Supplementary Table 2. Global cytokine table. Summary table for 57 cytokine values at four time points: initial rh-irAE onset, before ICI rechallenge, after rechallenge with TCZ prophylaxis, and at new irAE after rechallenge with TCZ prophylaxis: N, mean, min, max, percentiles (25th, 50th, 75th) and SD. The lower limit of detection is included in the table for all cytokines.

## Material and Methods

### Patient consent, ethical approval and sample collection

All study participants provided informed consent for the research use of their data through a process called "consentement général," which includes coded data handling to ensure confidentiality. For patients who did not actively object, enrollment was facilitated in accordance with Article 34 of the Swiss Federal Law on Human Research. This study was approved by the cantonal ethics committee, formally known as the Commission Cantonale d’Éthique de la Recherche sur l’Être Humain (CER-VD). Sample collection: All biological samples were collected during routine clinical practice. No specific interventions or modifications of standard care protocols were performed for the purposes of this study.

### Patient Cohort

#### Study design and population

This retrospective study, titled "Tocilizumab for Arthritis Prevention with Rh-irAEs (TAPIR study)," was conducted at the medical oncology and immunology services of the CHUV between 2020 and the end of 2023. It included a cohort of cancer patients treated with ICIs who developed ICI-AR (n=31). 4 patients declined the consent, 1 patient transferred to another hospital and only 26 were included in the final analysis, as displayed in the flowchart (**Supplemental Figure 1). The primary outcomes** were to assess the efficacy of TCZ in treating ICI-AR by evaluating the ACR 20/70 arthritis^25–27^ score and to determine its potential as a secondary prophylaxis during ICI re-challenge.

#### Treatment protocol

TCZ was administered at a dosage of 8 mg/kg biweekly (n=20). Concurrently, patients received low dose of prednisone (CS) at a dosing of 0.3 mg/kg (n=18), which was tapered down by 0.05 mg/kg weekly over a period of 6 weeks, until complete tapering. For those receiving secondary prophylactic treatment (TP) ((Rechallenge n=11, in 10 patients), TCZ was readministered at the same dosage of 8 mg/kg every two weeks concurrently with the ICI re-challenge treatment, without the introduction of CS. Another group of patients (Rechallenge n=5, in 3 patients) was rechallenged without TCZ (WP) prophylaxis as mentioned in the study design (**Supplemental Figure 2). Clinical follow-up**. Patients under TCZ treatment continued their routine clinical follow-up. The administration of TCZ did not influence oncological treatment decisions. All decisions regarding the ICI re-challenge were made by the referring oncologist. **Secondary outcomes**. ICI duration post rechallenge, reintroduction of CS >0.1mg/kg/day, ICI-RA flares and the objective response rate (ORR) and disease control rate (DCR)were monitored and recorded following the re-challenge with ICI therapy. Explanatory outcomes included biomarker identification that could predict response to TCZ treatment and prophylaxis (**Supplemental Figure 2**).

#### Immune profiling of serum biomarkers

As previously described^28^, serum concentrations of cytokines, chemokines, and growth factors were quantified in our study using the Luminex ProcartaPlex immunoassay panel. A comprehensive list of the biomarkers analyzed, their lower limits of detection (LLOD), and the distribution of values within our cohort is provided in Supplementary Table XX. Values recorded below or equal to the LLOD were adjusted to the LLOD to ensure analytical consistency. Our study included a comparative analysis of two different cohorts. In addition to the ICI-associated arthritis (ICI-AR) cohort of 26 patients, we collected baseline serum samples from a large cohort of cancer patients (n=164) prior to ICI treatment to determine pretreatment cytokine levels in a variety of tumor types. Cytokine levels were assessed longitudinally at several critical time points: at the onset of ICI-AR, at the time of ICI rechallenge, and throughout the course of TCZ prophylaxis treatment. This longitudinal approach allowed for a dynamic understanding of how cytokine profiles evolve in response to both disease progression and therapeutic interventions.

#### Statistical analysis

All statistical analysis in figures comparing cytokines were performed using GraphPad Prism 10.1.2 (GraphPad Software, Boston, Massachusetts USA).

## Discussion

The results of this study indicate that the early introduction of TCZ following the onset of ICI-RA, in conjunction with low-dose CS at 0.3 mg/kg, is highly effective for the treatment of ICI-RA, thereby eliminating the need for higher CS dosages. At 10 weeks, a 100% ACR70 response rate was achieved, with 81% of patients attaining CS-free remission after 24 weeks. These results are of particular significance as they demonstrate the rapid resolution of arthritis under TCZ, with a remarkable 70% symptom reduction within just 10 weeks of treatment across all participants. These outcomes represent a significant advancement over previous reports, as they establish a clear timeframe—24 weeks—for achieving steroid-free remission, a detail often omitted in earlier studies.

This strategy not only rapidly alleviates arthritis symptoms but also significantly improves the possibility of CS tapering. In particular, the duration to obtain CS<10mg was reduced by 24% compared to the CS group and by more than 50% compared to previous studies, which often reported treatment durations exceeding several months^1, 2, 4, 5^. The mean CS dose (mg) was reduced by 39% compared to the CS group. These results highlight TCZ’s potential to streamline and optimize arthritis treatment, reducing both the dose and the duration of CS treatment.

Another significant finding of this study was the absence of arthritis flares or the need for CS during rechallenge with ICIs when TCZ was used as a prophylactic agent. This outcome is of particular significance as it addresses an important challenge in the management of patients who have a history of ICI-RA. Typically, the recurrence of arthritis has constituted an obstacle to the continuation of potentially life-saving therapy. Our report underscores the efficacy of TCZ in preventing arthritis flares.

CS are frequently employed to address irAEs, but their immunosuppressive impact can diminish the advantages of ICIs^29–31^ and introduce complexities in long-term management due to their adverse effects. By reducing the necessity for CS in the treatment of ICI-RA, and by preventing the need for CS after ICI rechallenge, TCZ may help to preserve the integrity of the immune response initiated by ICIs, which is crucial for achieving optimal therapeutic outcomes. Moreover, as previously documented, IL-6 functions as a protumoral cytokine^32^. Consequently, the use of an anti-IL-6 strategy presents a dual benefit: firstly, it mitigates the adverse effects of CS and secondly, it enhances ICI efficacy by promoting Th1 polarization, thereby optimizing the therapeutic impact of ICIs^33–36^.

It is important to note that the use of TCZ as secondary prophylaxis significantly extended the duration of ICI administration, allowing re-administration for an average of 7.2 months compared to only 3.7 months without prophylaxis. In addition, TCZ prophylaxis significantly shortened the time to ICI rechallenge after initial discontinuation by a factor of 3.11, reducing the interval from 70 days to 22.5 days. At last follow-up, ongoing ICI use was 36% in the TCZ group (TP) versus 0% in the non-prophylaxis group (WP).

This is a retrospective study with a small number of patients and it does not allow statistical comparisons between patient groups. Importantly, the decision to rechallenge with ICI was made by oncologists based on evidence of disease progression or a perceived increased risk of progression. Consequently, these two groups of patients are distinct and cannot be directly compared. Nevertheless, we observed a sustained DCR in patients rechallenged with ICI under TCZ prophylaxis, even in patients presenting disease progression at rechallenge initiation. This finding is important because it demonstrates that TCZ prophylaxis may facilitate long-term use of ICIs and achievement of oncologic control, potentially improving overall survival (OS).

Our report suggests that TCZ can be effectively incorporated into the arthritis treatment regimen at an early stage and maintained without interruption of ICI therapy as a secondary prevention strategy. These results, among the first of their kind, set a new precedent for the management of ICI-RA and have the potential to significantly change current therapeutic paradigms by providing a more continuous and potentially more effective treatment pathway for patients undergoing ICI therapy.

This secondary prevention strategy, which involves the administration of TCZ following the onset of irAEs, contrasts with primary prevention, where TCZ is given preemptively to mitigate the frequency and severity of irAEs^37^. The determination of the most efficacious approach, whether primary or secondary prevention, requires further research. This necessity is further reinforced by the mounting body of evidence linking the occurrence of irAEs to the efficacy of ICI treatments^38–41^ and numerous studies that have reported elevated IL-6 levels during irAE episodes^42, 43^. Furthermore, the intricate network of cytokines may result in unfavorable outcomes when IL-6 signaling is blocked in the initial stages following the initiation of ICI therapy^44^. Accordingly, a more comprehensive investigation into the optimal timing for preventive strategies is of critical importance in this field.

In this study, we analyzed the levels of 57 cytokines at the onset of ICI-RA prior to the initiation of immunosuppressive (IS) treatment and compared these levels with baseline measurements in a cohort of cancer patients prior to ICI treatment (n=164). The increase in pro-inflammatory cytokines was limited to IL-6, CXCL9, CXCL10, and IL-RA. Other cytokine, chemokine and growth factor profiles remained stable. Notably, all patients with ICI-RA showed a consistent increase in IL-6 compared to baseline, without the concomitant increase in other inflammatory cytokines typically associated with inflammatory arthritis, such as IL-17^45^, IL-21^46^ and IL-22^47^. IL-6 levels continued to increase during TCZ treatment, as expected, and consistent with previous reports^11–13, 48, 49^. This distinct cytokine profile highlights the unique inflammatory pathway activated in ICI-RA and underscores the potential for therapeutic targeting of IL-6 in this context. Notably, while previous studies have reported the use of anti-IL-6 receptor therapy for the treatment of ICI-RA^17, 50^, they have not investigated longitudinal changes in cytokine levels. Our study fills this gap by assessing these variations, providing a deeper understanding of cytokine dynamics over the course of treatment and highlighting the potential importance of these changes in guiding therapeutic decisions. This approach not only validates the role of IL-6, but also suggests the utility of monitoring cytokine profiles as a component of personalized treatment strategies.

It is noteworthy that we observed an increase in CXCL9 levels during TCZ prophylaxis, similar to the levels observed at the onset of ICI-RA. This increase is clinically significant, as previously reported, as elevated serum CXCL9 during ICI-RA correlates with favorable oncological outcomes^51^. Furthermore, intratumoral increases in CXCL9, a chemokine essential for immune cell recruitment, have been associated with enhanced therapeutic responses in patients undergoing ICIs^52^. During TCZ prophylaxis, the stability of CXCL9 levels suggests that the inflammatory and immune-stimulatory milieu necessary for ICI efficacy is maintained. The non-reduction of CXCL9 levels suggest that, despite the immunosuppressive action of TCZ (which is primarily aimed at IL-6 blockade), the immune system’s ability to recruit T cells against the tumor remains active. This is of significant importance as it indicates that the prophylactic use of TCZ does not impede the primary anti-tumor immune response facilitated by ICIs. This is a significant concern when integrating any form of immunosuppression in cancer therapy. Monitoring CXCL9 levels may therefore prove to be a valuable biomarker for assessing the real-time efficacy of ICI therapy. By identifying and monitoring such cytokine surges, clinicians can better predict treatment responses and tailor therapeutic strategies, thereby optimizing patient outcomes. This approach capitalizes on the dual benefits of TCZ—managing arthritis and boosting ICI efficacy—through a deeper understanding of cytokine dynamics during treatment. The investigation of cytokine behavior in this context is of paramount importance and requires comprehensive investigation and validation to enhance our understanding of how immune modulation by TCZ impacts the success of cancer immunotherapy.

The observed increase in IL-6 and CXCL10 during new irAEs under TCZ prophylaxis suggest that IL-6 and CXCL10 warrant close monitoring, as they may signal the emergence of new irAEs. By allowing earlier detection, these biomarkers might prompt investigations and targeted interventions.

Although the initial results of our study are encouraging, several limitations must be acknowledged. First, the results are retrospective data derived from a single center with a relatively small cohort, which limits the generalizability of the results to different clinical settings. In addition, the study may lack diversity in patient demographics such as age, sex, ethnicity, and underlying health conditions, which may affect the applicability of the findings to broader populations. The potential retrospective design of the study also introduces inherent biases, such as selection bias and confounding factors, which require randomized controlled trials with large control arm without TCZ prophylaxis. Finally, more comprehensive studies are warranted to explore how modulation of immune responses by TCZ affects long-term oncologic efficacy.

In conclusion, this study suggests that TCZ is a viable strategy for both the treatment and prevention of ICI-RA upon ICI rechallenge. This dual efficacy of TCZ could significantly change the therapeutic landscape for patients undergoing ICI therapy who are at risk of, or have experienced, ICI-RA. By facilitating the broader use of ICIs in cancer patients, TCZ could ensure that a greater number of patients benefit from potent immunotherapy potentially improving patient outcomes by maintaining uninterrupted and effective ICI treatment.

## Data Availability

All data produced in the present study are available upon reasonable request to the authors

## Acknowledgements

This work was supported by the strategic plan of the CHUV. We would like to express our gratitude to all the patients who generously contributed their time and samples for this project. We thank Pr Gérard Waeber for his advice. Visual abstract was created with BioRender.com.

## Author contributions

MO conceived and designed the study. MO and PFP drafted the manuscript. MO, PFP and DD had full access to all data in the study and take responsibility for its integrity and accuracy.

DD, PFP and MO collected the data. PFP, DD and MO analyzed and interpreted the data. VJ and RB realized the cytokine data. SL, HB, NM, KA, JD, NF, LM, AS, KS and SP participated in the clinical treatments. PFP, DD and MO prepared the figures. SP. and GP participated in scientific discussion. The manuscript was reviewed and approved by all authors before submission.

## Data sharing statements

The datasets supporting the results of this study are not publicly available. Requests for access to the dataset will be granted upon reasonable request to the principal investigator. Study data will be managed, stored, shared, and archived according to CHUV standard operating procedures to ensure the continued quality, integrity, and utility of the data.

## References

1 Martins F, Sofiya L, Sykiotis GP et al. Adverse effects of immune-checkpoint inhibitors: epidemiology, management and surveillance. Nat Rev Clin Oncol 2019.

2 Calabrese LH, Calabrese C, Cappelli LC. Rheumatic immune-related adverse events from cancer immunotherapy. Nat Rev Rheumatol 2018; 14 (10): 569–579.

3 Belkhir R, Burel SL, Dunogeant L et al. Rheumatoid arthritis and polymyalgia rheumatica occurring after immune checkpoint inhibitor treatment. Ann Rheum Dis 2017; 76 (10): 1747–1750.

4 Braaten TJ, Brahmer JR, Forde PM et al. Immune checkpoint inhibitor-induced inflammatory arthritis persists after immunotherapy cessation. Ann Rheum Dis 2020; 79 (3): 332–338.

5 Ghosh N, Tiongson MD, Stewart C et al. Checkpoint Inhibitor-Associated Arthritis: A Systematic Review of Case Reports and Case Series. J Clin Rheumatol 2021; 27 (8): e317–e322.

6 Calabrese C, Cappelli LC, Kostine M et al. Polymyalgia rheumatica-like syndrome from checkpoint inhibitor therapy: case series and systematic review of the literature. RMD Open 2019; 5 (1): e000906.

7 Martin de Fremont G, Belkhir R, Henry J et al. Features of polymyalgia rheumatica-like syndrome after immune checkpoint inhibitor therapy. Ann Rheum Dis 2022; 81 (3): e52.

8 Jeurling S, Cappelli LC. Treatment of immune checkpoint inhibitor-induced inflammatory arthritis. Curr Opin Rheumatol 2020; 32 (3): 315–320.

9 Kim ST, Chu Y, Misoi M et al. Distinct molecular and immune hallmarks of inflammatory arthritis induced by immune checkpoint inhibitors for cancer therapy. Nat Commun 2022; 13 (1): 1970.

10 Wang R, Singaraju A, Marks KE et al. Clonally expanded CD38(hi) cytotoxic CD8 T cells define the T cell infiltrate in checkpoint inhibitor-associated arthritis. Sci Immunol 2023; 8 (85): eadd1591.

11 Doms J, Prior JO, Peters S et al. Tocilizumab for refractory severe immune checkpoint inhibitor-associated myocarditis. Ann Oncol 2020; 31 (9): 1273–1275.

12 Moi L, Bouchaab H, Mederos N et al. Personalized Cytokine-Directed Therapy With Tocilizumab for Refractory Immune Checkpoint Inhibitor-Related Cholangiohepatitis. J Thorac Oncol 2021; 16 (2): 318–326.

13 Ozdemir BC, Latifyan S, Perreau M et al. Cytokine-directed therapy with tocilizumab for immune checkpoint inhibitor-related hemophagocytic lymphohistiocytosis. Ann Oncol 2020; 31 (12): 1775–1778.

14 Kim ST, Tayar J, Trinh VA et al. Successful treatment of arthritis induced by checkpoint inhibitors with tocilizumab: a case series. Ann Rheum Dis 2017; 76 (12): 2061–2064.

15 Holmstroem RB, Nielsen OH, Jacobsen S et al. COLAR: open-label clinical study of IL-6 blockade with tocilizumab for the treatment of immune checkpoint inhibitor-induced colitis and arthritis. J Immunother Cancer 2022; 10 (9).

16 De La Fuente F, Belkhir R, Henry J et al. Use of a bDMARD or tsDMARD for the management of inflammatory arthritis under checkpoint inhibitors: an observational study. RMD Open 2022; 8 (2).

17 Fa’ak F, Buni M, Falohun A et al. Selective immune suppression using interleukin-6 receptor inhibitors for management of immune-related adverse events. J Immunother Cancer 2023; 11 (6).

18 Nakajima EC, Lipson EJ, Brahmer JR. Challenge of Rechallenge: When to Resume Immunotherapy Following an Immune-Related Adverse Event. J Clin Oncol 2019; 37 (30): 2714–2718.

19 Pollack MH, Betof A, Dearden H et al. Safety of resuming anti-PD-1 in patients with immune-related adverse events (irAEs) during combined anti-CTLA-4 and anti-PD1 in metastatic melanoma. Ann Oncol 2018; 29 (1): 250–255.

20 Simonaggio A, Michot JM, Voisin AL et al. Evaluation of Readministration of Immune Checkpoint Inhibitors After Immune-Related Adverse Events in Patients With Cancer. JAMA Oncol 2019; 5 (9): 1310–1317.

21 Dolladille C, Ederhy S, Sassier M et al. Immune Checkpoint Inhibitor Rechallenge After Immune-Related Adverse Events in Patients With Cancer. JAMA Oncol 2020; 6 (6): 865–871.

22 Haanen J, Ernstoff M, Wang Y et al. Rechallenge patients with immune checkpoint inhibitors following severe immune-related adverse events: review of the literature and suggested prophylactic strategy. J Immunother Cancer 2020; 8 (1).

23 Badran YR, Cohen JV, Brastianos PK et al. Concurrent therapy with immune checkpoint inhibitors and TNFalpha blockade in patients with gastrointestinal immune-related adverse events. J Immunother Cancer 2019; 7 (1): 226.

24 Badran YR, Zou F, Durbin SM et al. Concurrent immune checkpoint inhibition and selective immunosuppressive therapy in patients with immune-related enterocolitis. J Immunother Cancer 2023; 11 (6).

25 Felson DT, Anderson JJ, Boers M et al. American College of Rheumatology. Preliminary definition of improvement in rheumatoid arthritis. Arthritis Rheum 1995; 38 (6): 727–735.

26 van Gestel AM, Haagsma CJ, van Riel PL. Validation of rheumatoid arthritis improvement criteria that include simplified joint counts. Arthritis Rheum 1998; 41 (10): 1845–1850.

27 Prevoo ML, van’t Hof MA, Kuper HH et al. Modified disease activity scores that include twenty-eight-joint counts. Development and validation in a prospective longitudinal study of patients with rheumatoid arthritis. Arthritis Rheum 1995; 38 (1): 44–48.

28 Noto A, Joo V, Mancarella A et al. CXCL12 and CXCL13 Cytokine Serum Levels Are Associated with the Magnitude and the Quality of SARS-CoV-2 Humoral Responses. Viruses 2022; 14 (12).

29 Coutinho AE, Chapman KE. The anti-inflammatory and immunosuppressive effects of glucocorticoids, recent developments and mechanistic insights. Mol Cell Endocrinol 2011; 335 (1): 2–13.

30 Libert C, Dejager L. How steroids steer T cells. Cell Rep 2014; 7 (4): 938–939.

31 Giles AJ, Hutchinson MND, Sonnemann HM et al. Dexamethasone-induced immunosuppression: mechanisms and implications for immunotherapy. J Immunother Cancer 2018; 6 (1): 51.

32 Fisher DT, Appenheimer MM, Evans SS. The two faces of IL-6 in the tumor microenvironment. Semin Immunol 2014; 26 (1): 38–47.

33 Tsukamoto H, Fujieda K, Miyashita A et al. Combined Blockade of IL6 and PD-1/PD-L1 Signaling Abrogates Mutual Regulation of Their Immunosuppressive Effects in the Tumor Microenvironment. Cancer Res 2018; 78 (17): 5011–5022.

34 Mace TA, Shakya R, Pitarresi JR et al. IL-6 and PD-L1 antibody blockade combination therapy reduces tumour progression in murine models of pancreatic cancer. Gut 2018; 67 (2): 320–332.

35 Ohno Y, Toyoshima Y, Yurino H et al. Lack of interleukin-6 in the tumor microenvironment augments type-1 immunity and increases the efficacy of cancer immunotherapy. Cancer Sci 2017; 108 (10): 1959–1966.

36 Huseni MA, Wang L, Klementowicz JE et al. CD8(+) T cell-intrinsic IL-6 signaling promotes resistance to anti-PD-L1 immunotherapy. Cell Rep Med 2023; 4 (1): 100878.

37 J.S. Weber TM, O. Hamid, J. Mehnert, F.S. Hodi, S. Krishnarajapet, S. Malatyali, E. Buchbinder, J. Goldberg, R. Sullivan, M. Faries, I. Mehmi. Phase II trial of ipilimumab, nivolumab and tocilizumab for unresectable metastatic melanoma. Annals of Oncology 2021.

38 Shankar B, Zhang J, Naqash AR et al. Multisystem Immune-Related Adverse Events Associated With Immune Checkpoint Inhibitors for Treatment of Non-Small Cell Lung Cancer. JAMA Oncol 2020; 6 (12): 1952–1956.

39 Hu W, Wang G, Wang Y et al. Uncoupling Therapeutic Efficacy from Immune-Related Adverse Events in Immune Checkpoint Blockade. iScience 2020; 23 (10): 101580.

40 Hua C, Boussemart L, Mateus C et al. Association of Vitiligo With Tumor Response in Patients With Metastatic Melanoma Treated With Pembrolizumab. JAMA Dermatol 2016; 152 (1): 45–51.

41 Fujii T, Colen RR, Bilen MA et al. Incidence of immune-related adverse events and its association with treatment outcomes: the MD Anderson Cancer Center experience. Invest New Drugs 2018; 36 (4): 638–646.

42 Yu Y, Wang S, Su N et al. Increased Circulating Levels of CRP and IL-6 and Decreased Frequencies of T and B Lymphocyte Subsets Are Associated With Immune-Related Adverse Events During Combination Therapy With PD-1 Inhibitors for Liver Cancer. Front Oncol 2022; 12: 906824.

43 Zhang X, Lu X, Yu Y et al. Changes of IL-6 And IFN-gamma before and after the adverse events related to immune checkpoint inhibitors: A retrospective study. Medicine (Baltimore) 2022; 101 (46): e31761.

44 Wei F, Sasada T. Circulating cytokine signatures as a soluble biomarker of immune checkpoint inhibitor therapy in non-small-cell lung cancer. Genes Immun 2024; 25 (1): 89–91.

45 Gaffen SL. The role of interleukin-17 in the pathogenesis of rheumatoid arthritis. Curr Rheumatol Rep 2009; 11 (5): 365–370.

46 Agonia I, Couras J, Cunha A et al. IL-17, IL-21 and IL-22 polymorphisms in rheumatoid arthritis: A systematic review and meta-analysis. Cytokine 2020; 125: 154813.

47 Aarts J, Roeleveld DM, Helsen MM et al. Systemic overexpression of interleukin-22 induces the negative immune-regulator SOCS3 and potently reduces experimental arthritis in mice. Rheumatology (Oxford) 2021; 60 (4): 1974–1983.

48 Sun X, Fang C, Jin S et al. Serum IL-6 level trajectory for predicting the effectiveness and safety of tocilizumab in the treatment of refractory Takayasu arteritis. Eur J Intern Med 2024.

49 Horisberger A, La Rosa S, Zurcher JP et al. A severe case of refractory esophageal stenosis induced by nivolumab and responding to tocilizumab therapy. J Immunother Cancer 2018; 6 (1): 156.

50 Bass AR, Abdel-Wahab N, Reid PD et al. Comparative safety and effectiveness of TNF inhibitors, IL6 inhibitors and methotrexate for the treatment of immune checkpoint inhibitor-associated arthritis. Ann Rheum Dis 2023; 82 (7): 920-926.

51 Michel Obeid VJ, Douglas Daoudlarian, Robin Bartolini, Hasna Bouchaab, Sofiya Latifyan, Nuria Mederos, Karim Abdelhamid, Valérie Mosimann, Nabila Ferahta, Keyvan Shabafrouz, Giuseppe Pantaleo, and Solange Peters. High-dimensional longitudinal immune profiling uncovers a dual role of the CXCL9/CXCR3, CXCL13/CXCR5, and CCL11/CCL3 axis in the coupling of immune-related adverse events to immune checkpoint inhibitor response. Journal of Clinical Oncology 2024.

52 House IG, Savas P, Lai J et al. Macrophage-Derived CXCL9 and CXCL10 Are Required for Antitumor Immune Responses Following Immune Checkpoint Blockade. Clin Cancer Res 2020; 26 (2): 487–504.

